# Estimating vaccine-prevented disease outcomes when vaccination has only direct effects

**DOI:** 10.64898/2026.06.20.26356134

**Authors:** Fuhan Yang, Andrew Magee, Sinead E. Morris, Sarabeth Mathis, Ryan Wiegand, Danielle Iuliano, Matthew Biggerstaff, Scott W. Olesen

## Abstract

Vaccination can be a useful intervention for reducing infectious disease burden. Estimating numbers of vaccine-prevented health outcomes is one approach to quantifying the benefits of vaccination. Here we improve a method described by Foppa et al. (1) that assumes vaccination has only direct effects, that is, it cannot prevent infection or onward transmission of the disease. We rederive this method and derive an improved method that increases estimation accuracy with minimal additional analytical complexity. To evaluate the improved method, we simulated disease outbreaks and compared the accuracy of the two methods for estimating prevented disease outcomes. In 84% of simulations performed over a wide parameter space, the improved method had an equal or smaller estimation error compared to the original Foppa method, with 7.9-fold smaller mean error and 44-fold smaller standard deviation of errors. Our study improves a method for estimating prevented burden when assuming vaccination has only direct effects.

## Introduction

Vaccination is an important method to reduce the number of serious illnesses and deaths for a number of diseases (2). One way to quantify the benefits of vaccination is to estimate the number of disease outcomes, such as illnesses, hospitalizations, or deaths, that are prevented by vaccination. That is, the difference between the observed number of outcomes and the counterfactual number of outcomes that would have occurred in the absence of vaccination.

A variety of methods exist for estimating prevented burden. If vaccination is assumed to have indirect effects, that is, preventing infection or reducing onward transmission of the disease, then dynamical disease models (3–5) that can account for these effects are required. However, if vaccination is assumed to have only direct effects (2), that is, reducing the risk of disease progression among infected individuals but not preventing infection or reducing onward transmission, then simpler methods can be used (1,6–8).

An analytical method used by Foppa et al. (1), which assumes that vaccination has only direct effects, has been applied in estimating vaccine-prevented burden for influenza (1,9–12) and COVID-19 (13–16). This method, as commonly used, has two important limitations. First, its simplest implementation, which we refer to as “the original Foppa method,” treats vaccine effectiveness against disease progression as a fixed scalar and does not account for any delay between vaccination and vaccine effectiveness (2). Second, it makes optimistic and likely improbable assumptions about the relative timing of vaccination and disease outcomes.

Here, we rederive the original Foppa method, showing that it is one method in a more general class of methods that can incorporate time-varying vaccine effectiveness and different approaches to imputing the specific times of disease outcomes and vaccinations from the numbers of outcomes and vaccinations reported for discrete reporting periods. Next, we derive an “improved Foppa method” that (1) allows for a delay between vaccine administration and vaccine effectiveness and (2) makes more realistic assumptions about the timing of vaccinations and disease outcomes. We then simulate vaccination campaigns and disease outbreaks to generate disease outcome counts, vaccination counts, and a true number of prevented disease outcomes. We compare the accuracy of the original and improved Foppa methods for estimating the number of prevented outcomes over a wide parameter space.

## Methods

### Original Foppa method

We consider problems of this form: given (1) the observed number of health outcomes *y*_*k*_ in each reporting period *k* (e.g., a week or a month) in some population, (2) the proportion *u*_*k*_ of that population newly vaccinated in each reporting period, and (3) the vaccine effectiveness (VE) against that health outcome, estimate the counterfactual number *y*_*k*_* of health outcomes that would have occurred in the absence of vaccination (1). We define the “original Foppa method” as the one that makes the estimate:

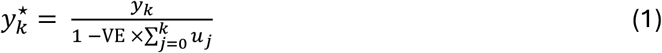

In other words, the number of observed outcomes *y*_*k*_ is the number of counterfactual outcomes *y*_*k*_*, reduced by the proportion of individuals who are effectively protected via vaccination. The estimated prevented burden is *y*_*k*_* - *y*_*k*_, that is, the difference between the estimated counterfactual number and the observed number of outcomes.

### Improved Foppa method

In Supplemental Information (S1.3), we show that the original method makes two important assumptions. First, it assumes that vaccination provides protection immediately upon administration (e.g., a very ill person might avoid hospitalization by being vaccinated just before they would be hospitalized). Second, it assumes that all vaccinations during each reporting period occur as an instantaneous impulse at the beginning of that period. These two assumptions are implausibly optimistic for most applications.

For purposes of this study, we define an “improved Foppa method” that has two improvements over the original Foppa method (Supplemental Information S1.4). First, we allow a nonzero “VE delay,” defined as the time between vaccine administration and the first moment when vaccination could prevent the health outcome. For example, say an individual must have been vaccinated at least *x* weeks before infection for vaccination to provide meaningful protection against hospitalization and that, in the absence of vaccination, they would become hospitalized *y* weeks after infection. In this example, the VE delay is *x* + *y* weeks: one must look back in time from the moment of the outcome and ask how long ago they would have needed to have been vaccinated in order to prevent that outcome. In the improved method, the VE delay is restricted to be an integer number *k*_min_ ≥ 0 of reporting periods.

Second, we assume that vaccinations and counterfactual health outcomes each occur at a constant rate during each reporting period. Under these assumptions, the estimated number of counterfactual outcomes in reporting period *k* is:

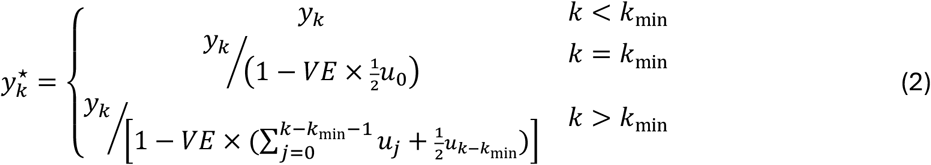

Note this is like the original Foppa method but with (1) vaccinations shifted forward in time by *k*_min_ reporting periods, indicating that vaccination only takes effect after the VE delay, and (2) only half the vaccinations from the most recent included period, due to the assumption about constant vaccination rates within each period.

### Simulation

To compare the accuracy of the original and improved Foppa methods, we perform stochastic simulations that track vaccinations and disease outcomes for each individual in the simulation.

First, the times of counterfactual health outcomes that would have occurred in the absence of vaccination are generated: a specified proportion of individuals are selected at random for counterfactual outcomes, then the times of those outcomes are selected at random from a normal distribution with specified mean and scale. Second, vaccination times are generated: a specified proportion of individuals are selected at random for vaccination, and the times of vaccine administration are selected at random from a specified distribution (also a normal distribution). Note that counterfactual outcome times and vaccination times are drawn independently. Third, health outcomes are determined: if a counterfactual outcome would have occurred more than the VE delay after that individual’s vaccination, the outcome is prevented with probability VE. Otherwise, the outcome occurs. Fourth, the numbers of vaccinations and outcomes per reporting period are summarized. Finally, the original and improved estimation methods are run using the vaccination and outcome time series, generating estimates of the number of counterfactual outcomes.

### Illustrative simulations

To confirm that the model broadly reproduces expected behavior, we ran simulations with a hand-picked set of parameters (Table 1) and visualized the results (Figure 1).

**Table 1.**
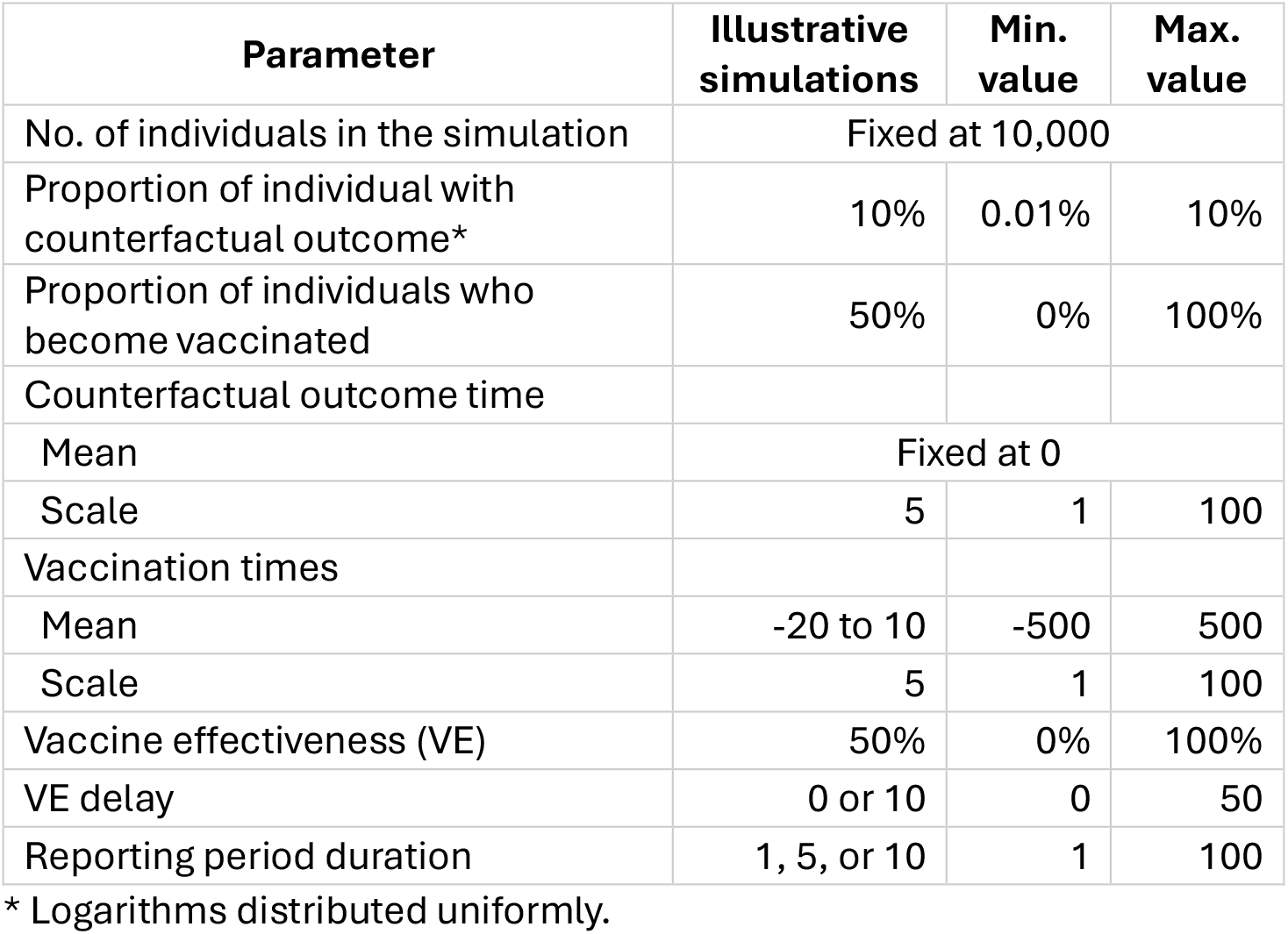
The ranges of parameters for the illustrative simulations and the methods comparison analysis. The outcome times, vaccination times, VE delays, and reporting period durations are all in arbitrary time units.

**Figure 1.**
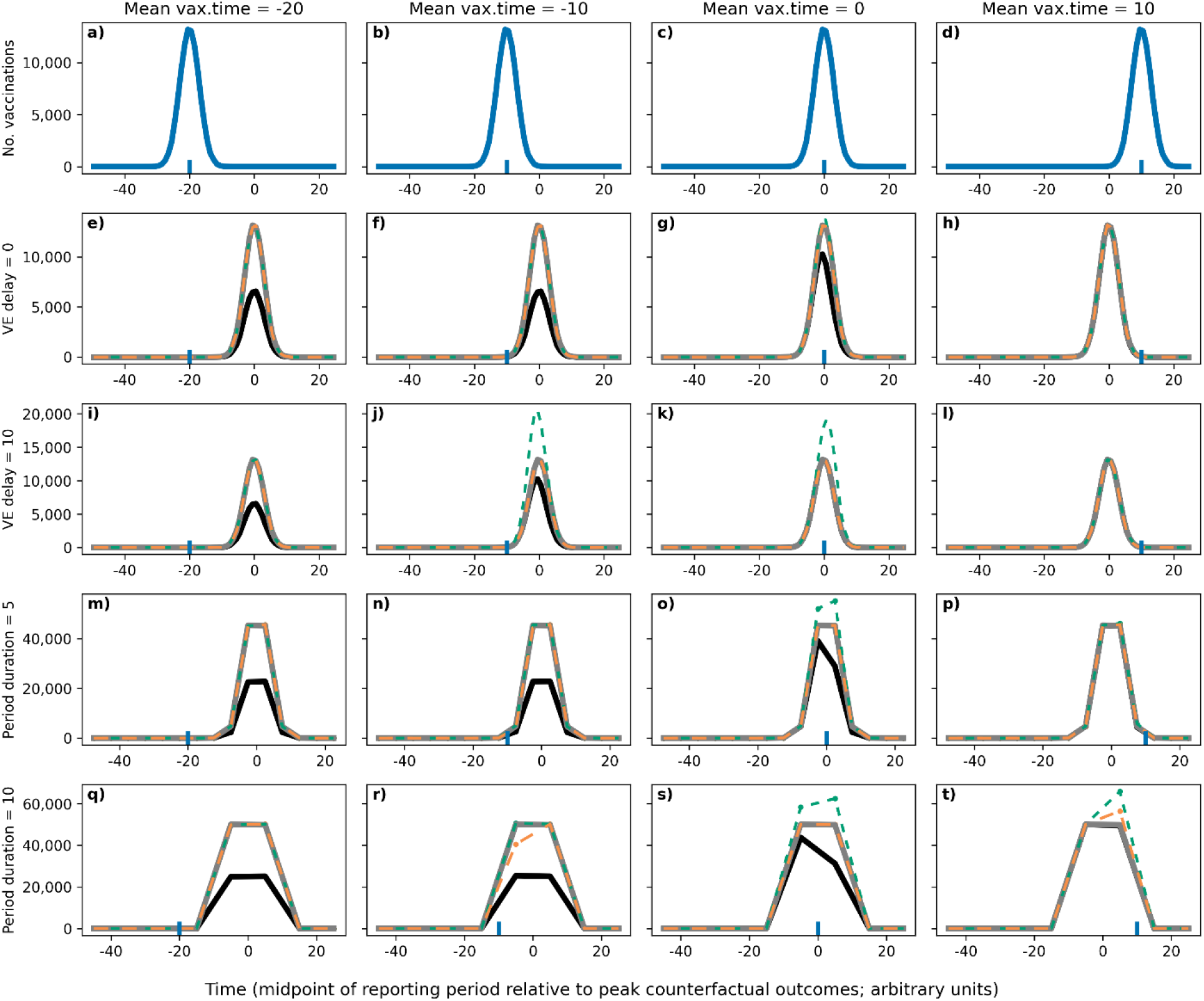
Illustration of vaccinations, counterfactual outcomes, outcomes, and estimated counterfactual outcomes. The first row (*a* through *d*) shows the distribution of vaccination times (blue lines) for four different mean vaccination times (blue ticks; -20 to +10). The second row (*e* through *h*) shows the counterfactual outcomes (gray line), outcomes (black line), and estimated counterfactual outcomes (original method, green; improved method, orange) for each mean vaccination time (column and blue tick) with no VE delay. (Note that the two estimation methods almost perfectly overlap the true values.) The third row (*i* through *l*) shows the same simulations but in the presence of a VE delay (10-time units). The fourth *(m* through *p)* and fifth rows *(q* through *t)* show the same simulations, with zero VE delay but with longer reporting periods (5- and 10-time units). Other parameters are as in Table 1.

### Method comparison

We performed 10,000 simulations over a wide parameter space using Latin hypercube sampling (Table 1). Parameters were each drawn uniformly between a minimum and maximum value, except for the proportion of individual with the counterfactual outcome, which was log-transformed. In the simulations, the reporting period and the true VE delay are both continuous-valued quantities; in the improved Foppa method, the VE delay is rounded to the nearest integer number of reporting periods. (For example, if the randomly drawn VE delay is 12.3 time units and the reporting period is 3.0 time units, then the VE delay is rounded to 4 reporting periods, that is, 12.0 time units.)

Simulations with no outcomes were discarded. The remaining simulations were analyzed by comparing the simulated number of counterfactual outcomes, which are defined as the “true” values, against the number of prevented outcomes estimated by the original and improved Foppa methods. The estimation error is the difference between the estimated number and the true simulated number of counterfactual outcomes. This value is equal to the error in the number of prevented outcomes, since the number of prevented outcomes is the difference between the estimated number of counterfactual outcomes and the fixed number of observed outcomes. Positive errors indicate overestimates (i.e., the vaccination campaign was estimated as more effective than it truly was). Negative errors indicate underestimates.

The statistical bias of each estimation method was computed as the mean error across simulations (with values closer to zero indicating less bias). The robustness of each estimation method was computed as the standard deviation of the errors across simulations.

### Correlation analysis

To evaluate the sensitivity of the relative performance of the two methods to specific parameterizations, the relative absolute error of the two methods for each simulation (i.e., the absolute error of the original method divided by the absolute error of the improved method) was computed, and those values were compared with each parameter’s value via a Kendall’s tau correlation.

### Code availability

Code to reproduce all results and figures are available at https://zenodo.org/records/20737466.

## Results

Figure 1 shows the illustrative simulations given the total number of individuals are 100,000, indicating some key patterns expected from the estimation methods. First, when vaccinations precede counterfactual outcomes, vaccinations provide protection, and the number of outcomes is smaller than the number of counterfactual outcomes (e.g., Figure 1e through 1g). Second, when vaccinations occur after the counterfactual outcomes, they do not provide protection, and numbers of outcomes and counterfactual outcomes are identical (e.g., Figure 1h). Third, in the presence of a VE delay, the original Foppa method tends to overestimate counterfactual outcomes (e.g., Figure 1j and 1k). Finally, in these illustrative scenarios, the improved method is usually more accurate than the original method (e.g., Figure 1o, 1s, and 1t) but not always (e.g., Figure 1r).

In the method comparison analysis, 9,889 of 10,000 simulations had at least 1 outcome and were included in the analysis (Figure 2). In 4,732 simulations (48%), the improved Foppa method had a lower absolute error than the original method. In 1,339 simulations (14%), the original method had a lower absolute error, and in 3,818 (39%), the two methods had the same error. Compared to the original method, the improved method was less biased (improved method mean error +0.2; original, +8.8) and more robust (improved method standard deviation of errors, 2.8; original, 22.1).

**Figure 2.**
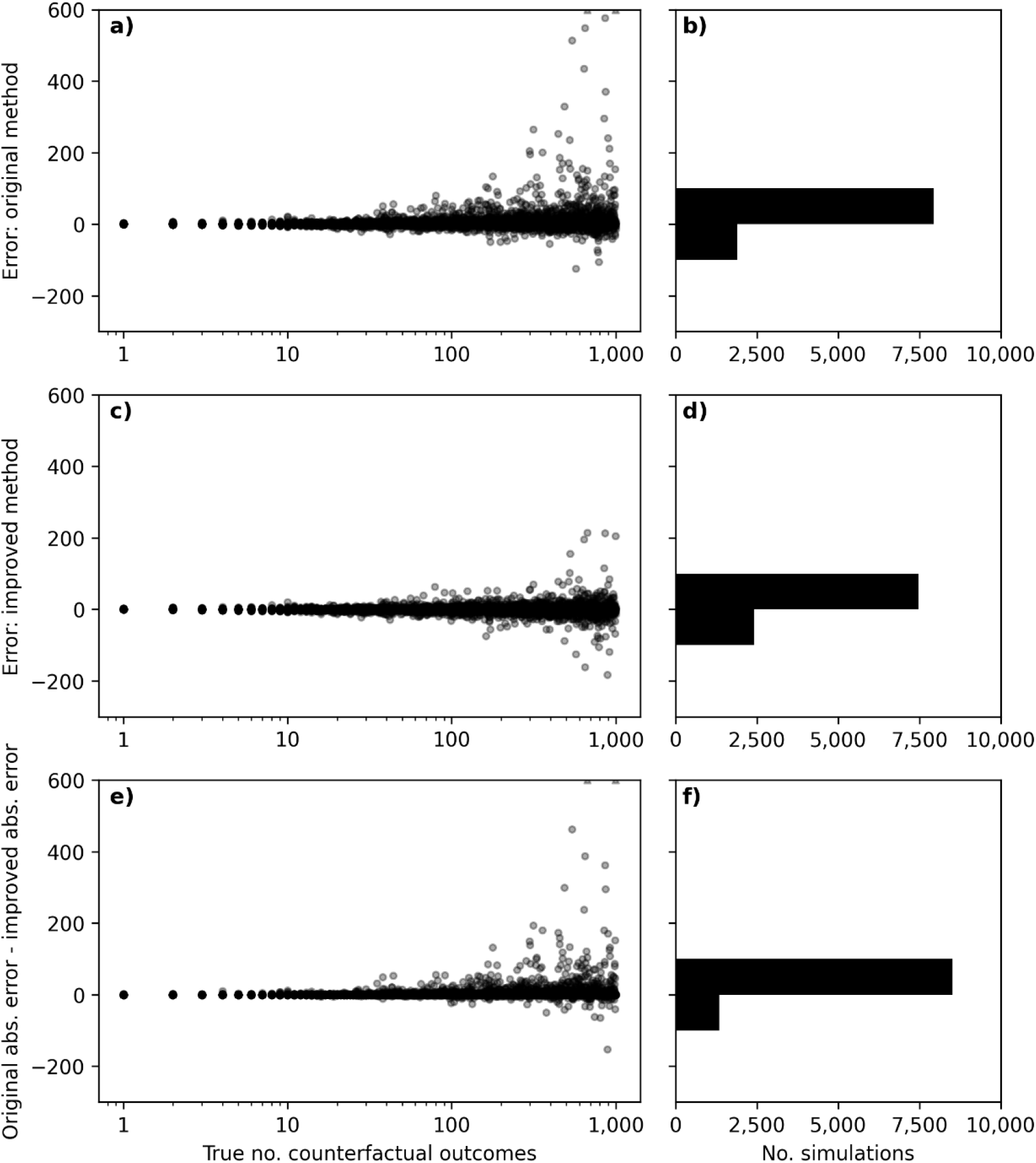
The performance of estimation methods. (a) Estimation errors of the original Foppa method (*y*-axis) for each simulation (point). Two outliers are shown at the upper *y*-axis limit. (c) Estimation errors for the improved method. (b, d) Histograms of the errors in *a* and *c*. (e) Difference in the absolute errors of the two estimation methods, where positive values indicate that the original method had a larger absolute error). (f) Histogram of values in *e*.

In the correlation analysis, only one parameter, mean vaccination time, had a strong (>0.2) correlation with the relative absolute error of the two methods (Figure S1, Table S1). The improved method performed especially well relative to the original method when the time of peak vaccinations came at or after the time of peak counterfactual outcomes (Table S2).

## Discussion

By re-deriving and generalizing the original Foppa method, we identified simple improvements and produced a method of comparable complexity that outperforms the original method over a wide parameter space. The improved method is less biased (i.e., expected value of its estimation error is closer to zero) and more robust (i.e., errors tend to be smaller).

The Foppa framework is flexible and can be adapted to different health outcomes and diseases. In this study, the improved method models VE as zero for some time since vaccination then a constant value, with no waning. The framework can accommodate other functional forms for VE, including arbitrary ramp up and waning (see Supplemental Information S1.4, Equation S4). The framework could even, with some extension, account for different counterfactual risks for the vaccinated and unvaccinated populations, for example, if the vaccinated population is at greater baseline risk of infection (Supplemental Information S1.5).

Despite their versatility, we note that Foppa framework methods have limitations. Most importantly, they assume that vaccination has only direct effects, which can underestimate prevented burden (19). Foppa framework methods appropriate only for use with vaccines that have limited effect on disease transmission. Second, Foppa framework methods are applicable for a single intervention, such as a single vaccination dose within a single season. Third, the structure of the improved Foppa method requires that the reporting period is shorter than the VE delay.

Overall, this study suggests that the improved Foppa method can be used for diseases with sufficiently well temporally resolved surveillance data and with a single-dose vaccine that does not substantially affect onward transmission.

## Data Availability

All code to reproduce all data are available at https://zenodo.org/records/20737466

## Acknowledgments

We thank Beau B. Bruce and Ivo Foppa for helpful comments.

## Disclaimer

The findings and conclusions in this report are those of the authors and do not necessarily represent the views of the Centers for Disease Control and Prevention.

