## Supplemental materials for "Estimating vaccine-prevented disease outcomes when vaccination has only direct effects"

### Supplementary Material: Estimating vaccine-prevented disease burden when vaccination has only direct effects

**S2 Figures and tables** **8**

### S1 Foppa framework

#### S1.1 Overview

Here we establish a rigorous mathematical foundation for the "original" and "improved" Foppa methods described in the main text.

Foppa framework methods take as input:

1. the number $n$ of individuals in a population,
2. the numbers $y_k$ of observed outcomes in that population in each reporting period $k$,
3. the proportions $u_k$ of that population newly vaccinated during each reporting period, and
4. vaccine effectiveness $\mathrm{VE}(\tau)$, as a function of time $\tau$ since vaccination,

and yield as output estimates $\hat{y}_k^\star$ of the numbers of outcomes that would have occurred in each period in the absence of vaccination. (We use the star superscript to designate quantities related to the no-vaccination counterfactual.) Note that the no-vaccination counterfactual refers to the absence of a vaccination *campaign*, not to an individual's vaccination status.

Assume that, in the absence of vaccination, each individual suffers outcomes as a Poisson process. Counterfactual outcomes occur at per-person rates $\lambda_k^\star$ that are constant within each period $k$, different between periods, and identical for all people. Let each period $k$ run from $t_k$ to $t_{k+1}$, with duration $T_k = t_{k+1} - t_k$. Then the number of counterfactual outcomes in each period is a random variable $Y_k^\star$, Poisson-distributed with mean $\mathbb{E}[Y_k^\star] = \lambda_k^\star n T_k$.

In the presence of vaccination, the outcome rate for each individual $i$ is:

$$\lambda(t, v_i) = \lambda_k^\star \left[1 - \mathrm{VE}(t - v_i)\right] \text{ for } t_k \leq t < t_{k+1}$$

where $v_i$ is the unobserved time of vaccination of person $i$. (By convention, to indicate that individual $i$ is never vaccinated, we set $v_i$ far in the future.) The number of outcomes in the presence of vaccination is a random variable $Y_k$ with mean:

$$\mathbb{E}[Y_k] = \sum_{i=1}^{n} \int_{t_k}^{t_{k+1}} \lambda(t, v_i)\, \mathrm{d}t = \lambda_k^\star \sum_{i=1}^{n} \int_{t_k}^{t_{k+1}} \left[1 - \mathrm{VE}(t - v_i)\right] \mathrm{d}t$$

We marginalize over the unobserved vaccination times $v_i$, treating them as independent and identically distributed random variables, with distribution $f_V$:

$$
\begin{aligned}
\mathbb{E}_V[\mathbb{E}Y_k] &= \lambda_k^\star \sum_{i=1}^{n} \int_{-\infty}^{\infty} \int_{t_k}^{t_{k+1}} [1 - \mathrm{VE}(t - v_i)]\, f_V(v_i)\, \mathrm{d}t\, \mathrm{d}v_i \\
&= \lambda_k^\star n \int_{-\infty}^{\infty} \int_{t_k}^{t_{k+1}} [1 - \mathrm{VE}(t - v)]\, f_V(v)\, \mathrm{d}t\, \mathrm{d}v \\
&= \lambda_k^\star n \cdot \left[ T_k - \int_{t_k}^{t_{k+1}} (\mathrm{VE} * f_V)(t)\, \mathrm{d}t \right]
\end{aligned}
$$

where the integral over $v$ is represented by the convolution $*$. For convenience, define:

$$
\alpha_k \equiv \frac{1}{T_k} \int_{t_k}^{t_{k+1}} (\mathrm{VE} * f_V)(t)\, \mathrm{d}t \tag{S1}
$$

so that $\mathbb{E}_V[\mathbb{E}Y_k] = \mathbb{E}[Y_k^\star](1 - \alpha_k)$.

Next, we generate method of moments estimators $\hat{y}_k^\star$ for $\mathbb{E}[Y_k^\star]$ that would equate the observed number $y_k$ of outcomes and the expected number $\mathbb{E}_V[\mathbb{E}Y_k]$ of those outcomes:

$$
y_k = \hat{y}_k^\star (1 - \alpha_k)
$$

In other words, the number of observed outcomes is equal to the estimated number of counterfactual outcomes, reduced by a factor $1 - \alpha_k$ that is related to vaccine effectiveness and coverage.

In Section S1.6 below, we show that the same equations result from more realistic assumptions about outcomes, that each person can suffer the outcome only once and that VE measures a reduction in hazard, not risk.

##### S1.2 Deriving specific Foppa framework methods

In principle, arbitrary $\mathrm{VE}(\tau)$ and $f_V(t)$ may be specified, and Equation S1 solved numerically. We refer to any such estimation method as a "Foppa framework method."

In practice, $\mathrm{VE}(\tau)$ is not known to such precision and is approximated by simpler functional forms, such as step functions. Furthermore, we typically have access only to the proportion $u_k$ of the population vaccinated in each time period, not to the full function $f_V(t)$.

Next, we show that simplifying assumptions about $\mathrm{VE}(\tau)$ and about $f_V(t)$ given the $u_k$ produce analytically tractable forms for $\alpha_k$.

##### S1.3 Original Foppa method

**VE** Under this method, we assume that vaccination is effective immediately upon administration and does not wane:

$$\mathrm{VE}(\tau) = \begin{cases} 0 & \tau < 0 \\ \mathrm{VE}_{\max} & \tau \geq 0 \end{cases}$$

where $\mathrm{VE}_{\max}$ is a fixed scalar. (In the main text, we use "VE" to refer to $\mathrm{VE}_{\max}$. Here we more carefully distinguish between time-varying $\mathrm{VE}(\tau)$ and the value $\mathrm{VE}_{\max}$ it takes on.)

**Vaccination rate as instantaneous impulses** This method makes the bounding assumption that all the vaccinations that occur during a period in fact occur as an instantaneous impulse at the beginning of that period:

$$f_V(t) = \sum_{k=-\infty}^{\infty} u_k T_k \cdot \delta(t - t_k) \tag{S2}$$

where $\delta(t)$ is the Dirac delta function. Then:

$$t \in [t_k, t_{k+1}) \implies (\mathrm{VE} * f_V)(t) = \mathrm{VE}_{\max} \cdot \sum_{\ell=-\infty}^{k} u_\ell T_\ell$$

This recovers the original Foppa method:

$$y_k = \hat{y}_k^{\star} \left( 1 - \mathrm{VE}_{\max} \cdot \sum_{\ell=-\infty}^{k} u_\ell \right)$$

where typically one assumes that $u_k = 0$ for $k < 0$ so that the sum begins at $\ell = 0$.

#### S1.4 Improved Foppa method

Assume that each reporting period $k$ is of equal duration $T$, and period 0 starts at $t = 0$, so that $t_k = kT$.

**VE** Assume that VE is a stepwise constant, with steps of duration equal to the reporting period $T$:

$$\mathrm{VE}(\tau) = \begin{cases} 0 & \tau \leq 0 \\ \mathrm{VE}_i & iT < \tau \leq (i+1)T \end{cases}$$

Note that VE varies only with time since vaccination, not with calendar time.

**Constant vaccination rates** Assume that the vaccination rate $f_V(t)$ is constant within each period:

$$f_V(t) = \frac{u_k}{T} \text{ for } kT \leq t < (k+1)T$$

Now derive:

$$\begin{aligned}
\alpha_k &= \frac{1}{T} \int_{kT}^{(k+1)T} (\mathrm{VE} * f_V)(t) \, \mathrm{d}t \\
&= \frac{1}{T} \int_{t=kT}^{(k+1)T} \int_{\tau=0}^{\infty} \mathrm{VE}(\tau) f_V(t-\tau) \, \mathrm{d}\tau \, \mathrm{d}t \\
&= \frac{1}{T} \int_{t=kT}^{(k+1)T} \sum_{i=0}^{\infty} \mathrm{VE}_i \cdot \int_{\tau=iT}^{(i+1)T} f_V(t-\tau) \, \mathrm{d}\tau \, \mathrm{d}t \\
&= \frac{1}{T} \sum_{i=0}^{\infty} \mathrm{VE}_i \cdot \int_{t=kT}^{(k+1)T} \{F_V(t-iT) - F_V(t-[i+1]T)\} \, \mathrm{d}t
\end{aligned}$$

Note that because $f_V(t)$ is piecewise constant, the cumulative coverage $F_V(t) \equiv \int_{-\infty}^{t} f_V(t') \, \mathrm{d}t'$ is piecewise linear. The integral $\int_a^b g(t) \, \mathrm{d}t$ of a function $g(t)$ that is linear over $(a, b)$ is $\frac{b-a}{2}[g(a) + g(b)]$. Thus, after some cancellations:

$$\alpha_k = \frac{1}{T} \sum_{i=0}^{\infty} \mathrm{VE}_i \cdot \frac{T}{2} \{F_V([k-i+1]T) - F_V([k-i-1]T)\}$$

Now note that $F_V(kT) = \sum_{\ell=-\infty}^{k-1} u_\ell$ so that:

$$\alpha_k = \frac{1}{2} \sum_{i=0}^{\infty} \mathrm{VE}_i \cdot (u_{k-i-1} + u_{k-i}) \tag{S3}$$

**VE step function** For purposes of the analyses in this study, we assume VE is a step function with no waning:

$$\mathrm{VE}_i = \begin{cases} 0 & i \leq k_{\mathrm{min}} \\ \mathrm{VE}_{\mathrm{max}} & i > k_{\mathrm{min}} \end{cases}$$

We refer to the time from vaccine administration to effectiveness $k_{\mathrm{min}}T$ as the "VE delay." Then:

$$\alpha_k = \frac{\mathrm{VE}_{\mathrm{max}}}{2} \cdot \{F_V([k - k_{\mathrm{min}}] \cdot T) + F_V([k - k_{\mathrm{min}} + 1] \cdot T)\} \tag{S4}$$

Assume also that there is some starting point $t = 0$ for vaccination, such that $u_k = 0$ for $k < 0$. Under these assumptions:

$$\alpha_k = \text{VE}_{\max} \cdot \begin{cases} 0 & k < k_{\min} \\ \frac{1}{2} u_0 & k = k_{\min} \\ \sum_{\ell=0}^{k-k_{\min}-1} u_\ell + \frac{1}{2} u_{k-k_{\min}} & k > k_{\min} \end{cases} \tag{S5}$$

which is referred to as the "improved Foppa method" in this study.

##### S1.5 Different counterfactual rates for vaccinated and unvaccinated

In the derivation above, we assumed common counterfactual rates $\lambda_k^\star$ for all individuals. Instead, separate individuals into two groups, $\mathcal{U}$ and $\mathcal{V}$, with separate rates and vaccination time distributions. Write the sizes of the groups as $|\mathcal{U}|$ and $|\mathcal{V}|$, with $|\mathcal{U}| + |\mathcal{V}| = n$.

For simplicity, consider a single time period and suppress the index $k$. Let individuals in group $\mathcal{U}$ have counterfactual rate $\lambda_{\mathcal{U}}^\star$ and similarly define $\lambda_{\mathcal{V}}^\star$. Assume that no individuals in $\mathcal{U}$ have been vaccinated as of this period so that $\lambda_{\mathcal{U}} = \lambda_{\mathcal{U}}^\star$.

Then the expected number of outcomes in this period in the absence of vaccination is $\hat{y}^\star = \left(|\mathcal{U}|\lambda_{\mathcal{U}}^\star + |\mathcal{V}|\lambda_{\mathcal{V}}^\star\right) T_k$, and the expected number in the presence of vaccination is:

$$\left[|\mathcal{U}|\lambda_{\mathcal{U}}^\star + |\mathcal{V}|\lambda_{\mathcal{V}}^\star \left(1 - \frac{n}{|\mathcal{V}|}\alpha\right)\right] T_k$$

Note the factor of $n/|\mathcal{V}|$, which accounts for the fact that $\alpha$ is defined with respect to the population-wide $f_V$, while the reduction in outcomes for $\mathcal{V}$ is related to the proportion of $\mathcal{V}$ that has been vaccinated.

Define the risk ratio $\text{RR} \equiv \lambda_{\mathcal{V}}^\star / \lambda_{\mathcal{U}}^\star$ so that the estimation equation is:

$$y = \hat{y}^\star \left(1 - \frac{n \cdot \text{RR}}{|\mathcal{U}| + |\mathcal{V}| \cdot \text{RR}} \alpha\right)$$

For higher risk ratios, more of the counterfactual outcomes are concentrated among the vaccinated, and a greater proportion of them are prevented by the same population-level vaccination corresponding to $\alpha$. Note that for $\text{RR} = 1$, this reduces to $y = \hat{y}^\star(1-\alpha)$, as in the prior derivation. For $\text{RR} = 0$, the vaccinated suffer none of the counterfactual outcomes, and $y = \hat{y}^\star$. For $\text{RR} \gg 1$, the vaccinated are the only ones at risk of the outcome, and $y \to \hat{y}^\star(1 - \alpha n/|\mathcal{V}|)$.

##### S1.6 Alternative derivation

Assume that each individual $i$ can suffer the outcome of interest only once and that all individuals have the same counterfactual hazard $r^\star(t)$ of suffering the outcome in the absence of vaccination.

The probability of having suffered the outcome by time $t$ in the absence of vaccination is:

$$p^\star(t) = 1 - \exp\left\{-\int_{-\infty}^{t} r^\star(s)\,\mathrm{d}s\right\}$$

Let the start and end times of each period be $t_k$ and $t_{k+1}$. Then the probability of suffering the outcome in period $k$ in the absence of vaccination is:

$$\begin{aligned}
\pi_k^\star &\equiv p^\star(t_{k+1}) - p^\star(t_k) \\
&= \left[1 - \exp\left\{-\int_{-\infty}^{t_{k+1}} r^\star(s)\,\mathrm{d}s\right\}\right] - \left[1 - \exp\left\{-\int_{-\infty}^{t_k} r^\star(s)\,\mathrm{d}s\right\}\right]
\end{aligned}$$

We linearize this result in the limit of small risk, i.e., $\int_{-\infty}^{t} r^\star(s)\,\mathrm{d}s \ll 1$. Note that, under a Taylor series approximation, $1 - e^{-x} = x + \mathcal{O}\left[x^2\right]$, so that:

$$\begin{aligned}
\pi_k^\star &= \int_{-\infty}^{t_{k+1}} r^\star(s)\,\mathrm{d}s - \int_{-\infty}^{t_k} r^\star(s)\,\mathrm{d}s + \mathcal{O}\left[\left(\int_{-\infty}^{t_{k+1}} r^\star(s)\,\mathrm{d}s\right)^2\right] \\
&\approx \int_{t_k}^{t_{k+1}} r^\star(s)\,\mathrm{d}s
\end{aligned}$$

Assume that the hazard is piecewise constant, such that $r^\star(t) = r_k^\star$ for $t_k \le t \le t_{k+1}$, and define the period durations $T_k = t_{k+1} - t_k$. Then:

$$\pi_k^\star \approx r_k^\star T_k$$

Vaccination reduces the hazard of the outcome. For an individual vaccinated at time $v$:

$$r(t; v) = r^\star(t)\,[1 - \mathrm{VE}(t - v)]$$

Analogous to the derivation above, the probability that an individual vaccinated at time $v$ suffers the outcome in period $k$ is:

$$\begin{aligned}
\pi_k(v) &\approx \int_{t_k}^{t_{k+1}} r(s; v)\,\mathrm{d}s \\
&= r_k^\star\left[T_k - \int_{t_k}^{t_{k+1}} \mathrm{VE}(t - v)\,\mathrm{d}t\right]
\end{aligned}$$

Next, we generate method of moments estimators $\hat{r}_k^\star$ for the counterfactual hazards, that is, we find the values for $r_k^\star$ that would equate $y_k$ and $\overline{y}_k \equiv \sum_{i=1}^{n} \pi_k(v_i)$, where $v_i$ are the times each individual is vaccinated. (By convention, to indicate that individual $i$ is never vaccinated, we set $v_i$ far in the future.)

Assume that the vaccination times $v_i$ are independent of one another, independent of counterfactual outcome times, and identically distributed according to some $f_V$. Marginalize over those times in the expectation:

$$\begin{aligned}\mathbb{E}_V[\overline{y}_k] &= \sum_{i=1}^{n} \mathbb{E}_V[\pi_k(v_i)] \\ &= n \int_{-\infty}^{\infty} \pi_k(v) f_V(v) \, \mathrm{d}v \\ &= n r_k^\star \left[ T_k - \int_{t_k}^{t_{k+1}} (\mathrm{VE} * f_V)(t) \, \mathrm{d}t \right]\end{aligned}$$

This reproduces the same method of moments estimator as in Section S1.1.

### S2 Figures and tables

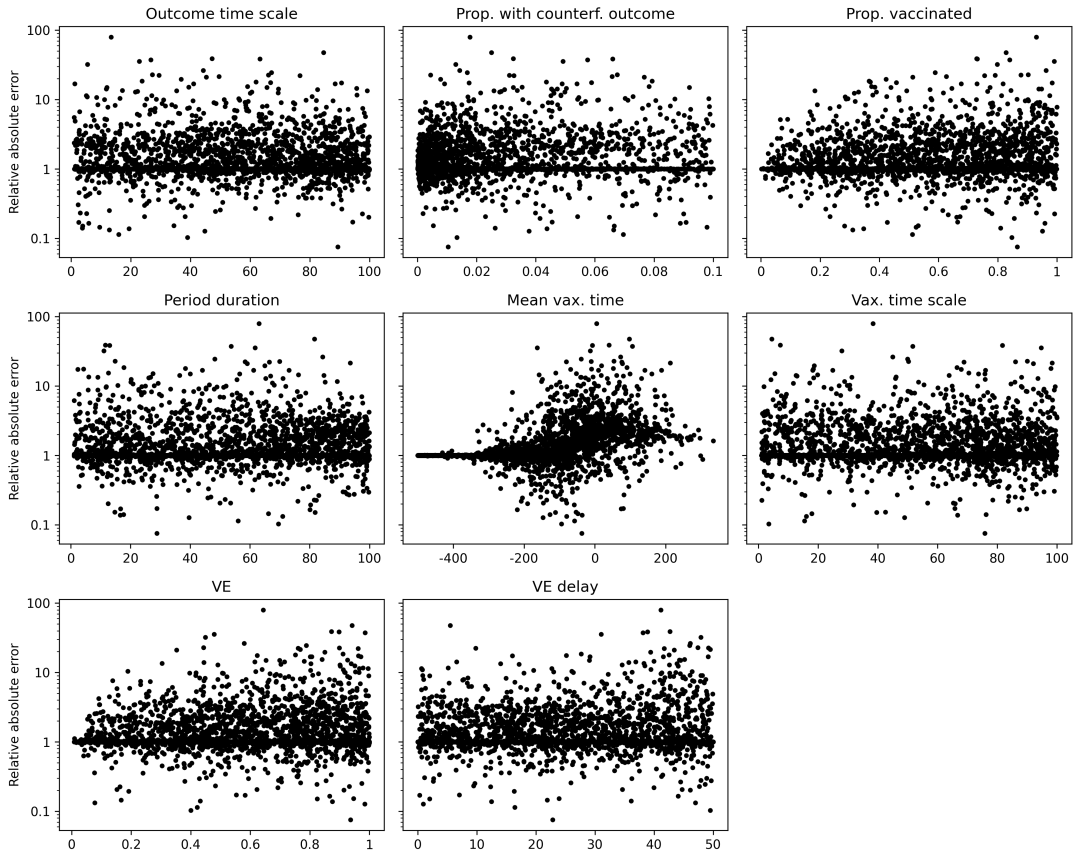

Figure S1: Correlation analysis. Relative absolute error ($y$-axis) for each simulation (points). Each facet orders the simulations by a different parameter ($x$-axis). Relative absolute errors greater than 1 indicate higher absolute error in the original method, relative to the improved method.

| Parameter | Correlation |
|---|---|
| Vaccination times - mean | 0.32 |
| Prop. of individuals with counterfactual outcome | 0.11 |
| VE | 0.05 |
| Reporting period duration | 0.05 |
| Prop. of individuals vaccinated | 0.04 |
| Counterfactual outcome times - scale | 0.03 |
| Vaccination times - scale | 0.03 |
| VE delay | 0.02 |

Table S1: Correlation analysis. Kendall's $\tau$ between relative absolute error and each parameter value across simulations.

| | Before | | After | |
|---|---|---|---|---|
| No. simulations | | | | |
| Total | 4,893 | | 4,996 | |
| Improved has lower error | 1,939 | (40%) | 2,793 | (56%) |
| Original has lower error | 1,137 | (23%) | 202 | (4%) |
| Equal errors | 1,817 | (40%) | 2,001 | (40%) |
| Mean error | | | | |
| Improved | −0.2 | | +0.6 | |
| Original | +2.7 | | +2.9 | |
| Mean absolute error | | | | |
| Improved | 4.4 | | 0.9 | |
| Original | 5.9 | | 2.9 | |

Table S2: Number of simulations (values) in which the time of peak vaccination comes before or after the time of peak counterfactual outcomes (columns).
